# Glucose Circadian Rhythm Disruption is Associated with Preeclampsia

**DOI:** 10.1101/2025.02.05.25321670

**Authors:** R Bravo, KH Lee, SA Nazeer, J Ashby Cornthwaite, B Sibai, M Fishel Bartal, C Pedroza

## Abstract

**Background:** There is emerging evidence of an association between circadian rhythms disruption and pregnancy complications. Preeclampsia is a leading cause of maternal death during pregnancy, and the role of circadian rhythms in predicting preeclampsia is not well understood.

**Objective:** Our aim was to determine whether glucose circadian rhythm disruption is associated with preeclampsia and can be used to predict this pregnancy disorder.

**Methods:** We analyzed a dataset of 92 recruited individuals with Continuous Glucose Monitoring (CGM) data recorded at 24.62 (sd = 4.97) weeks of gestational age. To study rhythmicity, we performed a cosinor analysis using the packages *cosinor* and *cosinor2*, and we calculated the non-parametric circadian rhythm variables using the *nparACT* package in R. Furthermore, we performed multiple component cosinor analysis to detect internal oscillations and identify glucose postprandial peaks using the package *CosinorPy* in Python.

**Results:** 71 participants (20 women with preeclampsia) had sufficient data for studying glucose circadian rhythmicity and performing cosinor analysis for multiple components to detect the postprandial peaks. We found that all the participants exhibited a significant circadian rhythm in their glucose oscillation. We developed a model including the time difference between the first postprandial peak and the last one, L5 start-time (time at which the five consecutive hours with the lowest average glucose levels start) and age that was predictive for preeclampsia incidence (AUC: 0.80 95%CI: 0.69-0.91, specificity= 0.88, sensitivity = 0.37). Patients diagnoses with preeclampsia from this model had a reduced amplitude (p < 0.05) and less robust (p < 0.05) glucose rhythmicity.

**Conclusion:** We conclude that evaluating glucose circadian rhythm during pregnancy may help to an earlier identification of preeclampsia.

## INTRODUCTION

Hypertensive disorders of pregnancy (HDP) can affect both the mother and the fetus, constituting the leading cause of maternal mortality^1^. Preeclampsia, a common pregnancy complication, is defined by a new onset of hypertension (> 140/90 mmHg) on 2 separate occasions after 20 weeks of gestation in previously normotensive individuals^2^. Often accompanied by proteinuria, preeclampsia may present with thrombocytopenia, impaired liver and/or renal functions, or pulmonary edema in the mother (Deepak, El-Balawi, and Harris,. It can also have significant effects on the fetus, including fetal growth restriction, preterm birth, or even intrauterine fetal death^3^. Current estimates indicate that the global incidence is about 4.6%. Although this percentage might vary depending on location, the incidence of preeclampsia is increasing worldwide^4^.

The clinical diagnosis of preeclampsia primarily depends on the assessment of blood pressure, urine protein or protein to creatinine ratio. However, this method has been reported as not sufficiently accurate or sensitive, leading to misdiagnosis and, consequently, delayed treatment^5^. Additionally, it has been reported that preeclampsia is closely related to changes in vasoconstriction, endothelial cells dysfunction, coagulation abnormalities, and immune dysfunction^6^. In this regard, most of the physiological variables associated with preeclampsia follow a 24-hour pattern. The pathophysiology of preeclampsia is not fully understood. Interestingly, preeclampsia is associated with gestational diabetes mellitus (GDM), diabetes type I (T1DM), and diabetes type II (T2DM), suggesting that glucose metabolism might be involved in the onset of preeclampsia^7,8^.

Circadian rhythms are those biological functions that follow a 24-hour pattern. Virtually every physiological function in the body shows a circadian oscillation that synchronizes internal clocks throughout the physiological systems, tissues, and cells. This synchronization is orchestrated by the suprachiasmatic nuclei, which, during nighttime, send signals to the pineal gland to release the indole melatonin and inform the body to perform nocturnal physiological functions^9^. Disruption of internal circadian rhythms, also known as chronodisruption, has been associated with many health impairments, as well as impaired reproductive function and hypertensive disorders of pregnancy like preeclampsia^10,11^. In healthy humans and healthy pregnant women, blood pressure follows a 24-hour pattern with higher values during the active phase and lower values during the sleep period. In fact, in healthy pregnant women, blood pressure dips 10-20 % less than daytime values. However, this oscillation is not that evident in patients with preeclampsia because the decrease during the night is less pronounced^12,13^. Moreover, circadian rhythms have been reported to be disrupted in women with preeclampsia. Actually, retrospective studies in humans have reported that sleep alterations or rotating shifts at work are related to a higher risk of hypertensive disorders of pregnancy, including preeclampsia^14– 16^. Furthermore, previous research has demonstrated that alterations in the molecular clock are associated with preeclampsia in both humans and mice^17^.

To date, the relationship between glucose circadian rhythms alterations and preeclampsia is limited, and more evidence is needed to assess whether abnormalities in circadian rhythms can predict preeclampsia. Therefore, our goal was to determine whether differences in glucose circadian rhythm, measured through Continuous Glucose Monitoring (CGM), may predict preeclampsia onset in pregnant individuals.

## METHODS

### Data

In this prospective observational study, 136 participants were initially enrolled between June 2020 and January 2022 for GDM screening at a single-level IV center. Eligibility criteria included age >18 years at ≤ 30 weeks of gestation. Exclusion criteria were known as type 1 or type 2 diabetes mellitus, history of bariatric surgery, or allergy to adhesive materials. For up to 10 days, participants wore a CGM device (G6 Pro CGM, Dexcom Inc, San Diego, CA) placed in blinded mode at the time of the 50-g GCT. During this time participants were asked to keep their usual diet and life schedule. Neither participants nor providers were able to access the CGM readings until the last recruited participant had delivered and all maternal and neonatal outcomes were collected. Further details of the study can be found in the previous publication^18^.

### Study outcomes

The primary outcome was the prevalence of preeclampsia depending on glucose rhythmic pattern. Secondary maternal outcomes were weight gain during pregnancy, induction of labor, HDPs, cesarean delivery, and postpartum complications, such as endometritis, wound infection, or wound dehiscence.

As neonatal outcomes we considered a composite adverse neonatal outcome defined as any of the following components: shoulder dystocia, large size for gestational age (LGA; defined as a birthweight of >90th percentile of the expected value according to gestational age using the nomogram by ^19^), respiratory distress (defined as needing at least 4 hours of respiratory support with supplemental oxygen, continuous positive airway pressure (CPAP), or ventilation at the first 24h of life), need for intravenous (IV) glucose therapy, and fetal or neonatal death. We also considered as neonatal outcomes GA at delivery, preterm delivery, 5-minute Apgar score of <7, NICU admission, small for gestational age (defined as birth weight of <10th percentile of the expected value according to GA), neonatal hypoglycemia (a blood glucose level of <40 mg/dL in the first hour of life or < 50 mg/dL after or requiring medical therapy), mechanical ventilation or CPAP, hyperbilirubinemia, hypocalcemia, and hospital length of stay.

### Chronobiological algorithms and metrics

To determine if any of the parameters obtained from glucose oscillation followed a circadian rhythm, we calculated chronobiological variables including MESOR, amplitude, acrophase, interdaily stability (IS), intradaily variability (IV), relative amplitude (RA) and percentage of rhythm using R software (version 4.1.1; R Core Team, 2013; R Foundation for Statistical Computing, Vienna, Austria). Glucose rhythmicity was calculated performing a cosinor analysis for every subject by fitting a sinusoidal function to the experimental variables with R packages *cosinor* and *cosinor2* (cosinor: http://github.com/sachsmc/cosinor, cosinor2: https://github.com/amutak/cosinor2). The mathematical expression is

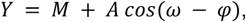

where M is the Midline Estimating Statistic Of Rhythm (MESOR), which is the estimate of central tendency of the distribution of values of an oscillating variable, A is the amplitude (difference between the peak and the MESOR) of the function, *ω* is the angular frequency equal to 2*π*/*T* with T being the period of the rhythm (i.e., T = 24h for circadian rhythms), and *φ*is the acrophase ^20,21^. Additionally, the model tests data, for every observation/volunteer, for circadian rhythmicity detecting a 24h pattern when p-value < 0.05 ^22^. Because the acrophases in the R packages mentioned above were expressed in radians, we converted them into hours for better comprehension. Moreover, to complement the cosinor models we calculated the Percent of Rhythm, which represented the percent of total variance accounted for by the fitted curve.

With the package *nparACT* ^23^ we additionally calculated the non-parametric circadian rhythm variables consisting of interdaily stability (IS), intradaily variability (IV) and relative amplitude (RA). The IS quantifies the invariability between the days, and provides an indication of the strength of coupling of the rhythm to a supposedly stable environmental zeitgeber. It was calculated as the ratio between the variance of the average 24-h pattern around the mean and the overall variance. Mathematically, it is expressed as follows:

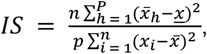

where ***n*** is the total number of data points, p is the number of data points per day, *<u>x</u>*_*h*_ are the hourly means, *<u>x</u>* is the mean of all data, and *x*_*i*_represents the individual data points collected.

The IV indicates the fragmentation of the rhythm, i.e., the frequency and extent of transitions between high and low values. It is calculated as the ratio of the mean square difference between successive hours (first derivative) and the mean square deviation from the grand mean (overall variance):

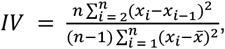

We also considered L5 and L5 start-time as variables which represent the five consecutive hours with the lowest average glucose levels and its time of onset. M10 and M10 start-time were also calculated, these variables express the 10 consecutive hours with the highest average glucose levels and its time of onset. RA is calculated by dividing amplitude by the sum of M10 and L5 ^24,25^.

Finally, to calculate the first and last postprandial glucose peaks and their timing for every participant we performed a multi-component cosinor analysis in Python using the package *cosinorPy* ^26^.

All of these chronobiological metrics obtained from fitting glucose values to a circadian rhythm were considered as potential predictors in the subsequent models.

### Statistical analysis

Continuous variables were summarized using means and their standard deviations and/or medians and interquartile ranges (IQRs). Categorical variables were depicted using frequencies and percentages. Maternal outcomes were compared between two groups using either the t-test or the Mann-Whitney where appropriate. For maternal categorical outcomes, comparisons between the two groups were made using the chi-square test or the Fisher exact test where appropriate. Neonatal outcomes were compared using generalized estimating equations (GEE) models to account for the correlation between twins in the study.

We constructed a receiver operating characteristic curve (ROC) to evaluate the diagnostic utility of each of the 11 chronobiological metrics for predicting preeclampsia incidence. We used the Youden index to choose thresholds for predicting preeclampsia incidence. To further assess performance, we calculated Brier scores. Considering that the sample size is relatively small, we added a bootstrap procedure in the analysis to obtain optimism-corrected performance metrics area under the curve (AUC), sensitivity, and specificity^27,28^.

Chronobiological metrics were then included in a multiple logistic regression to assess whether a combination of predictors would better predict preeclampsia incidence. Maternal age was also included as a covariate since it is well-known to be associated with preeclampsia incidence. Given the small sample size and number of events, we restricted the final model to have a total of 3 predictors. All analyses were performed in R software version 4.4.0., except the multi-component cosinor that was performed in Python 3.10.9.

## RESULTS

The data comprised 92 subjects who participated in the study between June 2020 and January 2022, with an interruption in recruitment due to the COVID-19 pandemic. In this study, we included 71 participants, as our calculations did not iterate for 21 patients due to insufficient days recorded to analyze the multiple components models. The assessed sample did not show differences with the original sample (Supplementary Table 1).

Among the 71 patients 19 were Hispanic (26.8%), 31 non-hispanic black (43.7 %) and 9 were non-hispanic white (12.7 %) (Table 1). In addition, 16 women were overweight (22.5%), 44 showed obesity (62%) and 11 were classified as normal weight (15.5%). Furthermore, 11 patients had chronic hypertension (15.5%), 19 reported gestational hypertension (26.8%), 20 patients reported preeclampsia (28.2%) and 11 preeclampsia with severe features (15.5%).

**Table 1.** Maternal characteristics.

| **Characteristic** | **N = 71<sup>1</sup>** |
| --- | --- |
| **Maternal age (y)** | 28 (23, 33) |
| **>= 35** | 14 (19.7%) |
| **Race and ethnicity** |  |
| Hispanic | 19 (26.8%) |
| Non-hispanic Black | 31 (43.7%) |
| Non-hispanic White | 9 (12.7%) |
| Other | 12 (16.9%) |
| **Nulliparous** | 17 (23.9%) |
| **BMI** | 32 (27, 36) |
| **BMI classification** |  |
| Normal weight | 11 (15.5%) |
| Obese | 44 (62.0%) |
| Overweight | 16 (22.5%) |
| **BMI >= 30** | 44 (62.0%) |
| **Smoking during pregnancy** | 5 (7.0%) |
| **Substance abuse during pregnancy** | 5 (7.0%) |
| **Insurance type** |  |
| Government | 44 (62.0%) |
| Private | 22 (31.0%) |
| Uninsured | 5 (7.0%) |
| **Twin pregnancy** | 6 (8.5%) |
| **Chronic hypertension** | 11 (15.5%) |
| **Gestational hypertension** | 19 (26.8%) |
| **Preeclampsia** | 20 (28.2%) |
| **Preeclampsia with severe features** | 11 (15.5%) |
| **Family history of diabetes mellitus** | 24 (33.8%) |
| **Completed a 1h GCT** | 68 (95.8%) |
| **1h CGT** | 113 (96, 129) |
| **1h CGT of >= 135 mg/dL** | 13 (19.1%) |
| **Completed a 3h 100 mg GCT** | 11 (15.5%) |
| **Time from 1h to 3h GCT (d)** | 22 (14, 36) |
| **Gestational diabetes mellitus diagnosed** | 1 (1.4%) |
| **Medication for diabetes mellitus during pregnancy** | 1 (1.4%) |

<sup>1</sup>Median (IQR); n (%)

### Chronobiological metrics to predict preeclampsia incidence

All participants exhibited rhythmicity in their glucose oscillations over a 24-hour period (see sample metrics in Supplementary Table 2). Table 2 presents the predictors showing that L5 start-time and the average time between the first and last postprandial peaks had the best performance in predicting preeclampsia based on their AUC and their Brier scores.

**Table 2.** Glucose circadian rhythm variables and receiver operating characteristic curve for each variable when predicting adverse neonatal composite outcome.

| Variable | Threshold | Specificity | Sensitivity | AUC | AUC: 95% CI<br>low | AUC: 95% CI<br>up | Brier<br>Score |
| --- | --- | --- | --- | --- | --- | --- | --- |
| **MESOR** | 111.60 | 0.71 | 0.52 | 0.54 | 0.39 | 0.69 | 0.194 |
| **Amplitude** | 8.40 | 0.40 | 0.90 | 0.64 | 0.51 | 0.78 | 0.190 |
| **Acrophase** | 16.18 | 0.65 | 0.52 | 0.54 | 0.38 | 0.69 | 0.198 |
| **Interdaily Stability** | 0.16 | 0.73 | 0.57 | 0.62 | 0.48 | 0.76 | 0.192 |
| **Intradaily Variability** | 0.90 | 0.47 | 0.62 | 0.47 | 0.33 | 0.62 | 0.197 |
| **Relative Amplitude** | 0.11 | 0.33 | 0.81 | 0.56 | 0.41 | 0.70 | 0.196 |
| **L5** | 94.47 | 0.31 | 0.81 | 0.50 | 0.35 | 0.64 | 0.196 |
| **M10** | 94.47 | 0.31 | 0.81 | 0.52 | 0.37 | 0.68 | 0.198 |
| **L5 start time** | 19.90 | 0.85 | 0.45 | 0.68 | 0.53 | 0.82 | 0.183 |
| **M10 start time** | 13.90 | 0.60 | 0.48 | 0.46 | 0.30 | 0.62 | 0.197 |
| **Percentage of Rhythm** | 0.11 | 0.42 | 0.90 | 0.65 | 0.52 | 0.78 | 0.185 |
| **Mean time between postprandial peaks** | 12.69 | 0.82 | 0.50 | 0.67 | 0.53 | 0.82 | 0.186 |
| **Average glucose postprandial peaks** | 115.30 | 0.55 | 0.65 | 0.55 | 0.40 | 0.70 | 0.195 |
| **Average glucose postprandial troughs** | 99.88 | 0.71 | 0.50 | 0.56 | 0.41 | 0.71 | 0.192 |

### Logistic regression results

We performed a logistic regression model including preeclampsia incidence as the dependent variable, while the time difference between the first and last postprandial glucose peaks and L5 start-time as predictors and controlling for age. This model (Table 3) showed that higher separation than 12.6 hours between first postprandial glucose peak and the last one had a positive effect on preeclampsia incidence (OR: 4.37, 95% CI: 1.24-16.6, p < 0.05), while later times for L5 start-time (OR: 1.10, 95% CI: 1.01-1.21, p<0.05) and age (OR: 1.11, 95% CI: 1.02-1.22, p<0.05) contributed positively to preeclampsia incidence. When tested for preeclampsia incidence, this model showed an AUC of 0.800 (95% CI: 0.691-0.910), 88 % specificity, and 36 sensitivity. In addition, after correction for optimism with internal validation through bootstrapping, we got the same values with an AUC of 0.802 (95% CI: 0.693 - 0.912), 88% specificity, 37% sensitivity, and Brier score of 0.156 (see Supplementary Table 3 for optimism metrics details).

**Table 3.** Logistic regression models for preeclampsia incidence.

| Characteristic | OR<sup>1</sup> | 95% CI<sup>1</sup> | p-value |
| --- | --- | --- | --- |
| **Time difference (h) between first postprandial glucose peak and the last postprandial glucose peak** |  |  |  |
| Average time < 12.69 h | — | — |  |
| Average time >= 12.69 h | 4.37 | 1.24, 16.6 | 0.024 |
| **L5 start-time** | 1.10 | 1.01, 1.21 | 0.038 |
| **Age** | 1.11 | 1.02, 1.22 | 0.014 |

<sup>1</sup>OR = Odds Ratio, CI = Confidence Interval

Furthermore, we studied differences between circadian variables between those predicted as preeclamptic pregnancies and non-preeclamptic by our mode (Supplementary Table 4). In this way, our model showed that those women predicted for preeclampsia had a reduced amplitude for glucose oscillation (p<0.001), but also a reduced robustness for rhythmicity (IS, p <0.05).

Based on this new model to predict preeclampsia, we did not observe any difference in maternal characteristics depending on the prediction of preeclampsia, except for a higher proportion of subjects above 35 years old (p = 0.05) (Supplementary Table 5). We did not observe any significant maternal adverse outcome (Table 4), but a higher proportion of preeclamptic patients was identified by our model (p = 0.07). When considering neonatal outcomes (Supplementary Table 6), our model showed that the group associated with preeclampsia had a reduced value for birth weight (p = 0.07).

**Table 4:** Maternal outcomes by our model to anticipate preeclampsia.

| Characteristic | mod12 <= 0.30<br>N = 48<sup>1</sup> | mod12 > 0.30<br>N = 23<sup>1</sup> | p-value<sup>2</sup> |
| --- | --- | --- | --- |
| **Weight gain during pregnancy (kg)** | 9.4 (9.6) | 12.8 (8.8) | 0.15 |
| **Induction of labor** | 21 / 46 (45.7%) | 11 / 23 (47.8%) | 0.86 |
| **Hypertensive disorders** | 21 / 47 (44.7%) | 12 / 23 (52.2%) | 0.56 |
| **Preeclampsia with severe features** | 4 / 47 (8.51%) | 6 / 23 (26.1%) | 0.070 |
| **Cesarean delivery** | 21 / 46 (45.7%) | 11 / 23 (47.8%) | 0.86 |
| **Primary cesarean delivery** | 10 / 46 (21.7%) | 5 / 23 (21.7%) | >0.99 |
| **EBL > 1,000 ml** | 3 / 48 (6.25%) | 1 / 23 (4.35%) | >0.99 |
| **Endometritis** | 2 / 46 (4.35%) | 0 / 23 (0%) | 0.55 |
| **Wound complications** | 1 / 46 (2.17%) | 0 / 23 (0%) | >0.99 |
| **Postpartum readmission** | 0 / 46 (0%) | 1 / 23 (4.35%) | 0.33 |

<sup>1</sup>Mean (SD); n / N (%)
<sup>2</sup>Welch Two Sample t-test; Pearson's Chi-squared test; Fisher's exact test

**Table 5.** Internal validation of the model to predict preeclampsia.

| AUC | Brier score | Specificity | Sensitivity | Accuracy | Balanced accuracy |
| --- | --- | --- | --- | --- | --- |
| 0.77 | 0.165 | 0.862 | 0.258 | 0.699 (0.577, 0.804) | 0.56 |

## DISCUSSION

Preeclampsia is considered to be one of the “great obstetrical syndromes” that causes 60,000 maternal deaths and more than 500,000 preterm births every year worldwide^29^. Classically, preeclampsia can be classified as early-onset preeclampsia and late-onset form^30,31^. The early-onset preeclampsia is defined as the form that develops before 34 weeks of gestation, and the late-onset form at or after 34 weeks of gestation. In the early onset form, preeclampsia has two stages: the first stage is a poor placentation, and the second stage is the maternal systemic disease. To date, there is growing evidence that pathophysiological functions involved in preeclampsia are normally regulated by the circadian system. In this way, multiple physiological functions following a 24-hour pattern appear dysregulated during preeclampsia^32^.

CGM is a non-invasive method that is gaining popularity to monitor subcutaneous interstitial glucose at a high-resolution level in a non-invasive way^33^. In the last years, it has also been used to investigate pregnancy complications, mostly related to diabetes in pregnancy ^18,34^. Currently, there is limited research using CGM focused on preeclampsia, and classical analyses like time in range, glucose variability, or mean differences have been computed to elucidate if CGM can be used to anticipate preeclampsia in T1DM due to their close association^35–37^. While Englund Ogge (2024) did not find any differences between preeclamptic women and non-preeclamptic women in patients with T1DM, Tiselko et al. (2022) reported a negative correlation between glucose variability and gestational age in T1DM with preeclampsia. Thus, these differences in glucose values to detect preeclampsia can only be applied to predict preeclampsia risk in T1DM. In our study, we included 71 pregnant individuals without diabetes to study whether a chronobiological approach could be used to predict preeclampsia. Studying the circadian pattern and even the timing of postprandial peaks showed differences in preeclamptic pregnancies compared to healthy pregnancies. We observed that individuals diagnosed later with preeclampsia had higher time differences between the first postprandial glucose peak and the last one, suggesting an extended eating schedule. Actually, those classified as preeclampsia gestations in our model showed more than a 12.6h difference between the first glucose postprandial peak and the last one. Experiments in rats have shown that time-restricted feeding (TRF) has been associated with reduced inflammation and pro-apoptotic markers found in preeclampsia^38^. Nighttime eating and later glucose peaks at night contribute to impaired glucose metabolism^39^. Actually, to control metabolic perturbations during gestation, TRF has been proposed as an effective way to align eating events with circadian physiology to optimize metabolism^40^. Studies suggesting that glucose peaks at night might have negative consequences on pregnancy outcomes have been focused on GDM^41^. Actually, obese women, a group with a higher risk of developing GDM and preeclampsia, among other pregnancy complications, have higher glucose peaks at night^42^.

Despite the limited research between glucose circadian rhythms and the incidence of preeclampsia, multiple investigations support the relationship between impaired glucose metabolism, proper rhythmic physiology, and the development of this hypertensive disease. The early onset form of preeclampsia is associated with fetal growth restriction, and it comes from poor placentation due to abnormal trophoblast invasion into the uterus. This abnormal invasion can be triggered by high glucose concentrations ^43,44^, provoking a shallow spiral artery remodeling during the first half of pregnancy^31^. This hyperglycemia linked to poor trophoblast migration has been linked to trophoblast inflammatory and antiangiogenic profile^45^. Recently, it has been reported that Bmal1 expression, a master circadian gene regulator, is needed to ensure trophoblast invasion, and when silenced, it represses trophoblast invasion via downregulation of metalloproteinases (TIMP3) ^46^. Furthermore, regarding rhythmicity in metabolism, it has been observed that placenta in women with preeclampsia express less melatonin receptor (both MT1 and MT2), while melatonin has promoting invasion effects on trophoblasts ^47^. Actually, MT1 receptor expression is maximal during the first trimester of pregnancy ^48^, suggesting trophoblasts require a rhythmic metabolism but also a rhythmic environment for proper functioning. Another process involved in these initial stages in preeclampsia is endothelial dysfunction, which leads to trophoblast invasion disorder that can affect maternal spiral artery remodeling ^44^. A phase I trial demonstrated that melatonin administration mitigated preeclampsia symptoms and improved endothelial function 49, reinforcing the idea that internal oscillating physiology and proper alignment with external zeitgebers might reduce preeclampsia incidence at the beginning of pregnancy ^50^. During the second stage, clinical symptoms are observed and abnormalities in nitric oxide (NO) metabolism are caused by the endothelial dysfunction started in the previous stage ^51^. Inhibition of NO metabolism increases platelet aggregation and inflammatory cell activation while impeding vasodilation ^52^. These symptoms, if untreated, will progress to high blood pressure, stroke, systemic organ failure, cerebral edema or seizures ^53,54^. It is known that melatonin from the maternal pineal gland increases during gestation in healthy pregnancies and shows lower levels in preeclamptic women. Actually, the deficiency in melatonin production during preeclamptic pregnancies is correlated with the severity of the disease ^55^. It is well-known that melatonin synchronizes internal circadian rhythms and is a hypertensive agent, promoting NO synthesis in endothelium. Although we did not measure the rhythmicity of any of these outcomes in our investigation, we observed a decreased amplitude in glucose oscillation in preeclamptic women, suggesting an impaired internal oscillation in other physiological functions. Furthermore, circadian rhythms in the body are interconnected ^56^, suggesting that the disruption observed in glucose rhythm could be present in other functions of the affected patients.

Finally, our study faced some limitations, like the sample size and, consequently, the number of women affected by preeclampsia. Actually, our algorithm to predict preeclampsia had a good sensitivity, but its ability to predict false negatives (specificity) was low and potential results may be masked by false positives. In addition, the lack of external validation is a common problem when approaching new perspectives. Moreover, evaluating sleep or even assessing chronotype could provide valuable information to understand the extent to which glucose circadian rhythm and sleep are interconnected during pregnancy. Finally, despite the limitations of our study, this research opens the possibility to investigate whether the glucose circadian rhythm of other circadian physiological functions may predict the incidence of preeclampsia earlier than current strategies.

## DATA AVAILABILITY

The datasets generated during and analyzed during the current study are available from the corresponding author on reasonable request pending application and approval by the research group.

