## Supplementary material for "Glucose Circadian Rhythm Disruption is Associated with Preeclampsia": Suplementary Tables

**Supplementary Table 1. Maternal characteristics comparison between current sample and original sample**

| **Characteristic** | **Circadian**, N = 71^1^ | **TAR**, N = 92^1^ | **p-value**^2^ |
| --- | --- | --- | --- |
| **Maternal age (y)** | 28.0 (23.0, 33.0) | 28.0 (23.0, 32.3) | 0.7 |
| **>= 35** | 14 (19.7%) | 16 (17.4%) | 0.7 |
| **Race and ethnicity** |  |  | >0.9 |
| Hispanic | 19 (26.8%) | 25 (27.2%) |  |
| Non-hispanic Black | 31 (43.7%) | 41 (44.6%) |  |
| Non-hispanic White | 9 (12.7%) | 11 (12.0%) |  |
| Other | 12 (16.9%) | 15 (16.3%) |  |
| **Nulliparous** | 17 (23.9%) | 25 (27.2%) | 0.6 |
| **BMI** | 32.4 (26.7, 36.4) | 31.2 (26.0, 36.2) | 0.5 |
| **BMI classification** |  |  | 0.9 |
| Normal weight | 11 (15.5%) | 16 (17.4%) |  |
| Obese | 44 (62.0%) | 53 (57.6%) |  |
| Overweight | 16 (22.5%) | 23 (25.0%) |  |
| **BMI >= 30** | 44 (62.0%) | 53 (57.6%) | 0.6 |
| **Smoking during pregnancy** | 5 (7.0%) | 5 (5.4%) | 0.7 |
| **Substance abuse during pregnancy** | 5 (7.0%) | 5 (5.4%) | 0.7 |
| **Insurance type** |  |  | 0.8 |
| Government | 44 (62.0%) | 60 (65.2%) |  |
| Private | 22 (31.0%) | 24 (26.1%) |  |
| Uninsured | 5 (7.0%) | 8 (8.7%) |  |
| **Twin pregnancy** | 6 (8.5%) | 3 (3.3%) | 0.2 |
| **Chronic hypertension** | 11 (15.5%) | 14 (15.2%) | >0.9 |
| **Family history of diabetes mellitus** | 24 (33.8%) | 29 (31.5%) | 0.8 |
| **Completed a 1h GCT** | 68 (95.8%) | 89 (96.7%) | >0.9 |
| **1h CGT** | 112.5 (95.8, 129.0) | 112.0 (95.0, 127.0) | 0.8 |
| **1h CGT of >= 135 mg/dL** | 13 (19.1%) | 17 (19.1%) | >0.9 |
| **Completed a 3h 100 mg GCT** | 11 (15.5%) | 14 (15.2%) | >0.9 |
| **Time from 1h to 3h GCT (d)** | 22.0 (13.5, 35.5) | 25.0 (18.5, 29.5) | 0.9 |
| **Gestational diabetes mellitus diagnosed** | 1 (1.4%) | 2 (2.2%) | >0.9 |
| **Medication for diabetes mellitus during pregnancy** | 1 (1.4%) | 1 (1.1%) | >0.9 |
| ^1^Median (IQR); n (%) | | | |
| ^2^Wilcoxon rank sum test; Pearson's Chi-squared test; Fisher's exact test | | | |

**Supplementary Table 2. Fitted cosinor model for glucose during 24h for multiple days in the studied sample**

| **Characteristic** | **N = 71**^1^ |
| --- | --- |
| **MESOR** | 105.7 (97.9, 112.7) |
| **Amplitude** | 6.7 (4.2, 8.8) |
| **Acrophase** | 14.6 (13.1, 18.0) |
| **Percentage of rhythm** | 0.1 (0.0, 0.1) |
| **Interdaily Stability (IS)** | 0.2 (0.2, 0.3) |
| **Intradaily Variability (IV)** | 0.9 (0.7, 1.0) |
| **Relative Amplitude (RA)** | 0.1 (0.1, 0.1) |
| **Average glucose value during the 10 consecutive hours with highest value (M10)** | 108.5 (101.2, 118.0) |
| **M10 start time (decimal time, h)** | 12.5 (6.6, 15.7) |
| **Average glucose value during the 5 consecutive hours with the lowest value (L5)** | 88.8 (81.4, 94.6) |
| **L5 start-time (decimal time, h)** | 12.7 (6.5, 19.9) |
| ^1^Median (IQR) | |

**Supplementary Table 3. Optimism correction for performance metrics obtained by bootstrap (1000 draws)**

| AUC | Brier score | Specificity | Sensitivity | Accuracy | Accuracy   (CI 95%) | Balanced Accuracy |
| --- | --- | --- | --- | --- | --- | --- |
| 0.0033 | -0.0018 | 0.0032 | 0.0022 | 0.0041 | 0.0053, 0.0025 | 0.0027 |

**Supplementary Table 4: Chronobiological variables depending on the model to predict preeclampsia (mod12 > 0.30 for preeclampsia prediction)**

| **Characteristic** | **mod12 <= 0.30**   N = 47^1^ | **mod12 > 0.30**   N = 23^1^ | **p-value**^2^ |
| --- | --- | --- | --- |
| **MESOR** | 105.69 (97.87, 112.58) | 105.73 (95.74, 115.43) | 0.54 |
| **Amplitude** | 7.20 (4.91, 9.30) | 4.88 (2.97, 6.07) | 0.002 |
| **Acrophase** | 14.49 (13.19, 17.28) | 17.19 (12.29, 19.48) | 0.30 |
| **Percentage of rhythm** | 0.10 (0.04, 0.13) | 0.04 (0.02, 0.05) | <0.001 |
| **Interdaily Stability** | 0.21 (0.16, 0.27) | 0.16 (0.11, 0.21) | 0.022 |
| **Intradaily Variability** | 0.83 (0.69, 0.96) | 0.95 (0.80, 1.17) | 0.069 |
| **Relative Amplitude** | 0.10 (0.08, 0.12) | 0.09 (0.07, 0.12) | 0.68 |
| ^1^Median (Q1, Q3) | | | |
| ^2^Welch Two Sample t-test | | | |

**Supplementary Table 5:  Maternal characteristics of the study population by our model to predict preeclampsia (mod12 > 0.30 for preeclampsia prediction)**

| **Characteristic** | **mod12 <= 0.30**   N = 48^1^ | **mod12 > 0.30**   N = 23^1^ | **p-value**^2^ |
| --- | --- | --- | --- |
| **Maternal age (y)** | 27.0 (23.0, 31.5) | 29.0 (22.0, 35.0) | 0.36 |
| **>= 35** | 6 (12.5%) | 8 (34.8%) | 0.052 |
| **Race and ethnicity** |  |  | 0.98 |
| Hispanic | 13 (27.1%) | 6 (26.1%) |  |
| Non-hispanic Black | 20 (41.7%) | 11 (47.8%) |  |
| Non-hispanic White | 6 (12.5%) | 3 (13.0%) |  |
| Other | 9 (18.8%) | 3 (13.0%) |  |
| **Nulliparous** | 13 (27.1%) | 3 (13.0%) | 0.19 |
| **BMI** | 32.6 (27.0, 36.1) | 32.8 (26.3, 43.1) | 0.84 |
| **BMI classification** |  |  | 0.57 |
| Normal weight | 7 (14.6%) | 3 (13.0%) |  |
| Obese | 32 (66.7%) | 13 (56.5%) |  |
| Overweight | 9 (18.8%) | 7 (30.4%) |  |
| **BMI >= 30** | 32 (66.7%) | 13 (56.5%) | 0.41 |
| **Smoking during pregnancy** | 4 (8.33%) | 1 (4.35%) | >0.99 |
| **Substance abuse during pregnancy** | 4 (8.33%) | 1 (4.35%) | >0.99 |
| **Insurance type** |  |  | 0.60 |
| Government | 31 (64.6%) | 13 (56.5%) |  |
| Private | 13 (27.1%) | 9 (39.1%) |  |
| Uninsured | 4 (8.33%) | 1 (4.35%) |  |
| **Twin pregnancy** | 4 (8.33%) | 2 (8.70%) | >0.99 |
| **Chronic hypertension** | 8 (16.7%) | 4 (17.4%) | >0.99 |
| **Family history of diabetes mellitus** | 13 (27.1%) | 10 (43.5%) | 0.45 |
| **Completed a 1h GCT** | 46 (95.8%) | 22 (95.7%) | >0.99 |
| **1h CGT** | 112.5 (95.0, 127.0) | 113.0 (96.0, 129.0) | 0.65 |
| **1h CGT of >= 135 mg/dL** | 7 (15.2%) | 5 (22.7%) | 0.50 |
| **Completed a 3h 100 mg GCT** | 7 (14.6%) | 3 (13.0%) | >0.99 |
| **Gestational diabetes mellitus diagnosed** | 1 (2.08%) | 0 (0%) | >0.99 |
| **Medication for diabetes mellitus during pregnancy** | 1 (2.08%) | 0 (0%) | >0.99 |
| ^1^Median (Q1, Q3); n (%) | | | |
| ^2^Wilcoxon rank sum test; Fisher's exact test; Pearson's Chi-squared test | | | |

**Supplementary Table 6: Neonatal outcomes by our model to anticipate preeclampsia
(mod12 > 0.30 for preeclampsia prediction)**

| **Characteristic** | **mod12 <= 0.30**   N = 48^1^ | **mod12 > 0.30**   N = 23^1^ | **p-value**^2^ |
| --- | --- | --- | --- |
| **Composite neonatal outcomes** | 14 (29.8%) | 8 (34.8%) | 0.7 |
| **Shoulder dystocia or birth injury** | 1 (2.17%) | 0 (0%) | >0.9 |
| **Large for gestational age** | 4 (8.70%) | 2 (8.70%) | >0.9 |
| **Need for IV glucose** | 7 (15.2%) | 4 (17.4%) | >0.9 |
| **Respiratory distress** | 5 (10.9%) | 5 (21.7%) | 0.3 |
| **Fetal or neonatal death** | 1 (2.13%) | 1 (4.35%) | >0.9 |
| **GA at delivery** | 38.3 (37.0, 39.6) | 38.1 (36.3, 39.1) | 0.4 |
| **Preterm delivery (<37 wk)** | 10 (21.3%) | 7 (30.4%) | 0.4 |
| **Indicated preterm delivery** | 8 (17.0%) | 7 (30.4%) | 0.2 |
| **Birth weight** | 3,210.0 (2,890.0, 3,500.0) | 2,920.0 (2,610.0, 3,290.0) | 0.073 |
| **Lowest neonatal glucose** | 42.0 (30.5, 58.5) | 50.0 (44.0, 60.0) | 0.2 |
| **5-min Apgar score of <7** | 1 (2.17%) | 1 (4.55%) | 0.5 |
| **Small for gestational age** | 5 (10.9%) | 1 (4.35%) | 0.7 |
| **NICU admission** | 18 (39.1%) | 10 (43.5%) | 0.7 |
| **Hypoglycemia** | 11 (23.9%) | 4 (17.4%) | 0.8 |
| **Mechanical ventilation** | 2 (4.35%) | 1 (4.35%) | >0.9 |
| **Need for CPAP** | 5 (10.9%) | 5 (21.7%) | 0.3 |
| **Hyperbilirubinemia** | 7 (15.2%) | 5 (21.7%) | 0.5 |
| **Hypocalcemia** | 2 (4.35%) | 1 (4.35%) | >0.9 |
| **Length of hospital stay** | 2.0 (2.0, 4.0) | 2.0 (2.0, 3.0) | 0.7 |
| ^1^n (%); Median (Q1, Q3) | | | |
| ^2^Pearson's Chi-squared test; Fisher's exact test; Wilcoxon rank sum test | | | |
